# Mendelian Randomization Unveils a Causal Relationship Between Inflammation, Metabolism, and Systemic Sclerosis

**DOI:** 10.1101/2025.04.02.25325153

**Authors:** Yuanyuan Zeng, Wen Zeng, Zengyong Li, Xiaolan Yang, Jie Pan, Ling Lei

## Abstract

**Background:** Inflammatory processes and metabolic activity significantly influence the beginning and course of systemic sclerosis (SSc). This study aims to explore the genetic basis for the impact of inflammation and metabolism on susceptibility to SSc.

**Methods:** We used three types of exposure: 91 inflammatory proteins (14,824 participants), 731 immune cell traits (3,757 Sardinians), 1,400 blood metabolites (8,299 Europeans), with SSc as an outcome (680 cases, 399,355 controls). A two-sample multivariate bivariate Mendelian randomization (MR) was conducted to investigate the causal relationship between inflammation, metabolism, and SSc and explore the interaction of inflammatory proteins, immune cells, and metabolites in SSc.

**Results:** MR analysis identified potential causal associations of four inflammatory proteins and fourteen metabolites with SSc and a significant causal association of two immune cells with SSc. Among them, HLA DR on CD14- CD16+ monocyte and HLA DR on CD33dim HLA DR+ CD11b+ significantly reduced the risk of SSc. Pleiotropy and heterogeneity were not observed. None showed bidirectional causality in reverse MR analysis. Multivariate MR results showed independent causal effects of two inflammatory proteins, one immune cell, and three metabolites on SSc; three were risk factors (hepatocyte growth factor, stem cell growth factor, and 5-hydroxyindole sulfate levels); three were protective factors (HLA DR on CD14- CD16+ monocyte, Homoarginine levels and Tetradecadienoate (14:2) levels).

**Conclusions:** These findings reveal causal relationships and interactions between four inflammatory proteins, fifteen immune cell traits, and fourteen metabolites and SSc and its development, offering fresh perspectives on the mechanisms underlying SSc and guiding the choice of treatment approaches.

## 1. Introduction

Systemic sclerosis (SSc) is a complex and multifaceted autoimmune disease characterized by microvascular damage, excessive collagen deposition, and immune regulatory imbalance, leading to fibrosis and vascular lesions in the skin and various internal organs [1].According to the most recent study, the prevalence of SSc varies from 3.1 to 144.5 per 100,000 people, while the global incidence falls between 0.2 to 7.5 per 100,000 individuals per year [2]. Over 50 percent of SSc patient deaths are closely linked to the illness, and between 2000 and 2011, there was a gradual increase in this percentage [3]. SSc presents a diverse clinical spectrum involving multiple organs, making early diagnosis, especially in the absence of skin lesions, particularly challenging. The limited therapeutic options contribute to a significant clinical, social, and economic burden for patients with SSc, negatively impacting their quality of life and prognosis [4]. Consequently, the identification of appropriate biomarkers for diagnosis or targeted therapy is of paramount importance.

The etiology of systemic SSc remains incompletely understood, with current consensus suggesting that genetic susceptibility, environmental factors, and immune system dysregulation collectively contribute to its pathogenesis [5, 6]. Prior research has established the participation of immune cells [7], B cells [8], macrophages [9], interleukin-6 (IL-6) [10], IL-12 [11], and other inflammatory factors in the pathogenesis of SSc, underscoring the importance of inflammation and immune responses in this disease. Moreover, with advances in metabolomics technology, research has revealed significant differences in metabolic profiles between SSc patients and healthy individuals, particularly affecting the tricarboxylic acid cycle, fatty acid β-oxidation, and processes associated with amino acids. These metabolic disturbances may be linked to inflammation, vascular damage, fibrosis, and dysbiosis of the intestinal flora [12]. Recent evidence suggests that the inflammatory process and metabolism are essential factors in the development and course of SSc [13, 14]. These studies provide the foundation for future research by offering fresh perspectives on the pathophysiology of SSc while recognizing possible treatment strategies.

Nevertheless, previous research has mainly consisted of observational studies limited by the number of samples and confounding variables, which in some cases led to controversial conclusions. For instance, a different study discovered no appreciable variations in serum IL-17 and IL-6 levels between SSc patients and controls, despite Rositsa Karalilova et al. [15]reporting considerably higher levels of IL-6 in SSc compared to healthy comparators [16]. Research has identified dysfunctional circulating Treg cells in SSc patients, attributed to impaired secretion of IL-10 and TGFβ within these cells [17]. However, most studies have observed increased Treg cell numbers in SSc patients [18]. T_H17_ cells are prominently enriched in SSc patients and positively correlated with disease activity [19], though their precise role in SSc fibrosis remains uncertain [20]. Despite many observational studies yielding relatively consistent results, they often provide only correlative conclusions, thereby complicating establishing causal relationships and bidirectional associations with SSc. In contrast to conventional observational research, Mendelian randomization (MR) is a novel epidemiological analysis method that evaluates the causal links between exposure factors and diseases by using single nucleotide polymorphisms (SNPs) as instrumental variables (IVs). Genetic variation is randomly assigned at conception and follows chronological sequencing. Therefore, molecular variation effectively manages confounding variables and reverses causation problems [21]. This work uses a two-sample multivariable bidirectional MR study design that integrates large datasets to examine the genetic correlation of 91 inflammatory proteins, 731 immune cell characteristics, and 1,400 blood metabolites on SSc. The goals are to find new therapeutic targets and offer SSc patients more potent treatment plans.

## 2. Materials and methods

### 2.1 Study Design

We utilized two-sample MR to explore the genetic basis for the impact of inflammation and metabolism on susceptibility to SSc and identify potential treatment avenues. In this analysis, the validity of IVs hinges on their adherence to three key criteria: (1) a robust association with the Exposure, (2) no linkage to confounding variables, and (3) an exclusive relationship with SSc mediated solely through the exposure. We have examined the causal associations between 1400 blood metabolites, 731 immune cell features, and 91 inflammatory proteins and SSc in independent two-sample multivariate bidirectional MR (Fig 1). Because the data were freely accessible, there was no need for an ethics committee review.

**Fig 1.**
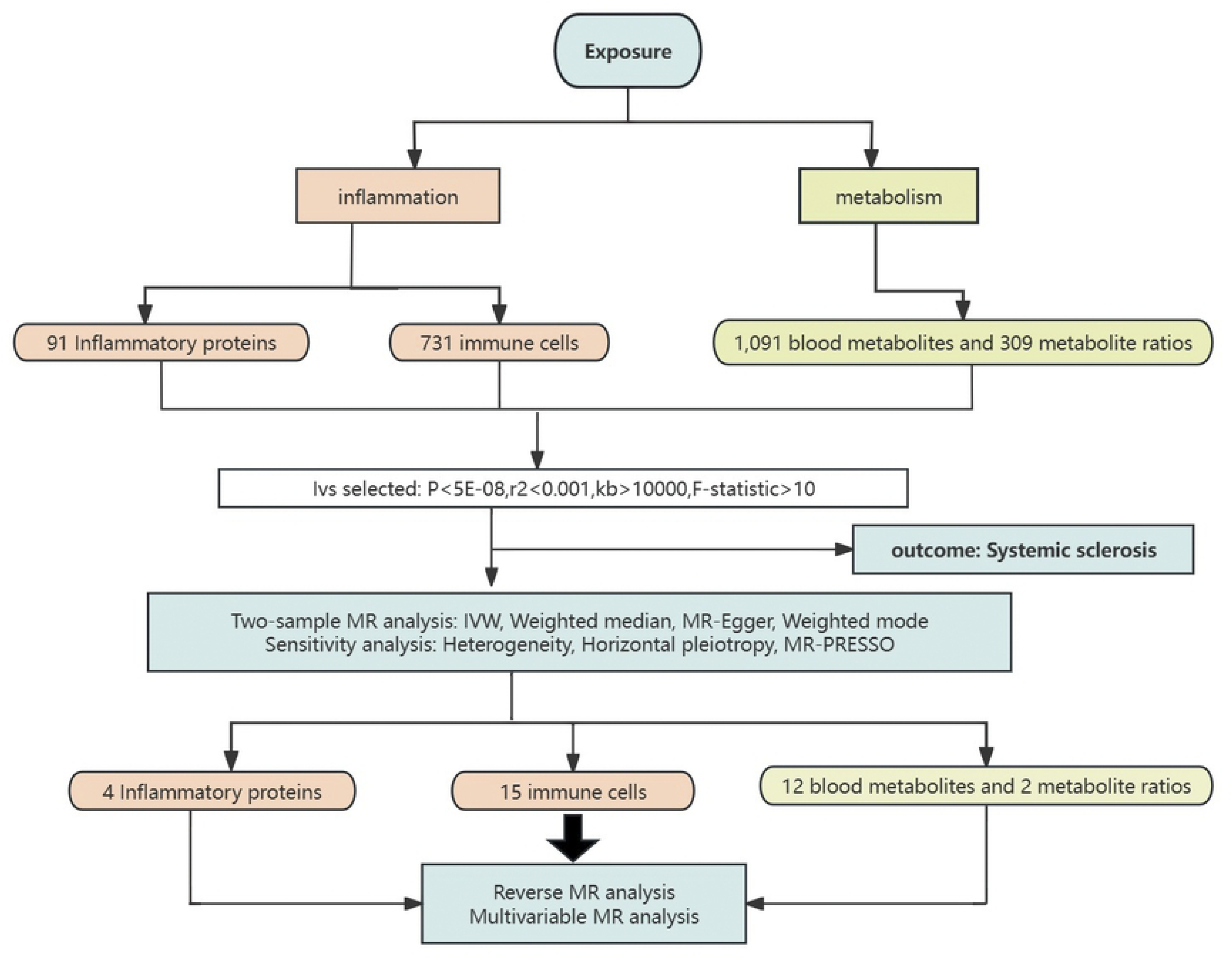

### 2.2 Data Source

The data were divided into two main components: exposure and outcome. Information about SSc cases was obtained from the latest and most comprehensive version, R10 of the FinnGen consortium (https://www.finngen.fi/en/access_results), which included 680 cases and 399,355 controls. The two most important and noticeable categories in the exposure data were inflammation and metabolism. The metabolism evaluation utilized 1400 blood metabolites extracted from the genome-wide association studies (GWAS) Catalog (GCST90199621-902010209). This catalog represents the most extensive genetic study of blood metabolite loci, involving 8,299 individuals of European descent. The study identified genetic associations of 690 metabolites at 248 loci and 143 metabolic ratios at 69 loci [22], providing crucial insights into the relationships between genetic variations and metabolites. The inflammation analysis was divided into 91 inflammatory proteins and 731 immune cell traits. Using the Olink Target platform, 14,824 participants were assessed for 91 plasma proteins, identifying 180 genome-wide protein quantitative trait loci (59 cis-effects, 121 trans-effects) [23]. In the immune cell data analysis, a cohort of 3,757 Sardinian residents was studied, examining over 22 million SNPs associated with 731 immune cell features and identifying 53 novel loci involved in cellular regulation across various molecules and mechanisms [24]. Additional details can be found in Supplementary Table S1.

### 2.3 Genetic Instrumental Variable Selection

We used strict genetic IVs in this part to ensure our investigation was reliable and robust. We identified SNPs significantly associated with the phenotype using a stringent threshold (*P* < 5 × 10^−8^). All GWAS datasets in this study provide P-values for SNP-exposure associations, as detailed in Supplementary Tables S2-S4 displaying pval.exposure. To exclude SNPs in high linkage disequilibrium (LD) (r^2^ < 0.001, kb > 10,000), we used quality control criteria. We combined datasets and assessed each one’s consistency with exposure and outcome. We also adjusted palindromic SNPs with unclear alleles to guarantee consistency using allele frequency data. Ultimately, we calculated F-statistics and established a cutoff point (F > 10) to eliminate possibly weak independent variables and mitigate bias between exposure and instrumental variables. Detailed SNP information was presented in Supplementary Tables S2-S4.

### 2.4 MR Analysis and Sensitivity Analysis

We examined the R 4.1.2 software’s “Two-Sample MR” and “MRPRESSO” packages for this investigation. The odds ratio (OR) and its 95% confidence interval (CI) were primarily calculated using the Inverse Variance Weighted (IVW) technique in order to assess any potential causal relationships between exposure and outcome [25]. Furthermore, we supplemented our analysis with MR-Egger regression [26], weighted median [27], and weighted mode [28]. Sensitivity tests were carried out to validate the validity and dependability of the MR analysis. SNP heterogeneity was measured and evaluated using Cochran’s Q [29]. A random-effects model was applied if *p* < 0.05, indicating heterogeneity; otherwise, a fixed-effects model was used. We employed the MR-Egger and MR pleiotropy residual and outlier (MRPRESSO) methods to evaluate horizontal pleiotropy [30]. To guarantee the validity of the conclusions, exposure data demonstrating horizontal pleiotropy were also eliminated. We applied the Benjamini-Hochberg method to address multiple testing issues, integrating false discovery rate (FDR) [31]. An association between the exposure and SSc was possible if the exposure’s initial p-value was less than 0.05, but its FDR-corrected p-value was higher than 0.05. Nevertheless, if the original and FDR-corrected p-values were less than 0.05, it was deemed to have a significant causal connection with SSc.

### 2.5 Reverse Causality Detection

Following the same selection criteria mentioned above, we conducted a bidirectional Mendelian randomization analysis with SSc as the exposure and the identified positives (inflammation and metabolites) as the outcome to explore potential reverse causal relationships. The subsequent stages were identical to those in the MR forward process.

### 2.6 Multivariable MR

In practice, specific genetic variants are associated with multiple risk factors, a phenomenon known as pleiotropy [32]. In these situations, numerous potentially interacting exposures might be included in multivariable Mendelian randomization (MVMR) to adjust for the interaction of genetic variations among exposures. Univariate MR evaluates the total effect of an exposure on an outcome, whereas MVMR assesses the direct effect of each exposure on the outcome, independent of any other exposures. Given the interacting associations between inflammation and metabolites, we conducted MVMR in this study on the identified inflammatory proteins, immune cells, and metabolites to adjust for their interactions.

## 3 Results

### 3.1 Bidirectional MR Analysis of Inflammatory Proteins on SSc

After stringent control of IV quality, the MR study identified a total of 73 inflammatory proteins. All included SNPs had F-statistics greater than 10, indicating strong IV performance (Supplementary Table S2). Subsequent MR analysis revealed significant estimates from IVW (*p* < 0.05). Consistent directions and magnitudes were observed across IVW, MR-Egger, and weighted median estimations. Ultimately, four inflammatory proteins were identified as potentially causally affecting SSc, including Oncostatin-M (OR 2.65, 95% CI: 1.41-4.99, *p =* 0.002), Hepatocyte growth factor (OR 3.96, 95% CI: 1.59-9.85, *p* = 0.003), Macrophage inflammatory protein 1a (OR 1.46, 95% CI: 1.08-1.98, *p* = 0.013), and Stem cell factor (OR 1.48, 95% CI: 1.08-2.03, *p* = 0.015) (Fig 2). MR-PRESSO analysis of these positive exposures revealed no heterogeneous SNPs. Cochran’s Q test (*p* > 0.05) and MR-Egger intercept test (*p* > 0.05) provided strong evidence of no heterogeneity or pleiotropy (Supplementary Table S8), suggesting these four proteins are candidates for further analysis. After applying FDR correction, the p-values for these proteins were all higher than 0.05 (Supplementary Table S5), indicating a potential causal association with SSc. In order to minimize the possibility of reverse causality, we used SSc as the exposure and the four inflammatory proteins as the outcomes of reverse MR analysis. The results revealed no evidence of bidirectional causality (*p* > 0.05) (Supplementary Table S11.1).

**Fig 2.**
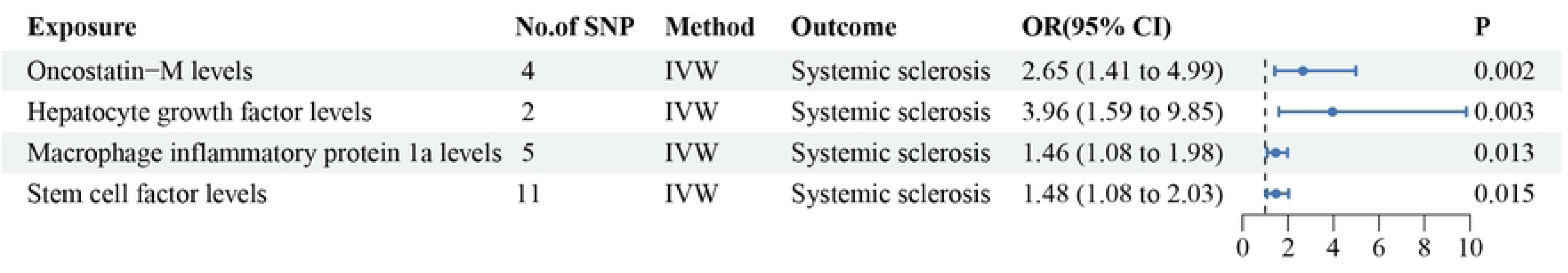

### 3.2 Bidirectional MR Analysis of Immune Cell Traits on SSc

After stringent control of IV quality, we ultimately captured 603 immune cell phenotypes. All included SNPs had an F statistic more significant than 10 (Supplementary Table S3). Subsequent MR analysis showed significant estimates from IVW (*p* < 0.05). Consistent directions and magnitudes were observed across IVW, MR-Egger, and weighted median estimations. We ultimately identified fifteen immune cells that may causally influence SSc. Among them, seven decreasing the risk of SSc: HLA DR on CD14- CD16+ monocyte (OR 0.69, 95% CI: 0.60-0.81, *p* < 0.001), HLA DR on CD33dim HLA DR+ CD11b+ (OR 0.70, 95% CI: 0.58-0.84, *p* < 0.001), CD28 on secreting CD4 regulatory T cell (OR 0.63, 95% CI: 0.45-0.87, *p* = 0.006), CD28 on CD39+ activated CD4 regulatory T cell (OR 0.56, 95% CI: 0.38-0.85, *p* = 0.006), CD4+ CD8dim T cell %lymphocyte (OR 0.65, 95% CI: 0.44-0.95, *p* = 0.028), CD45RA+ CD8+ T cell %CD8+ T cell (OR 0.92, 95% CI: 0.85-0.99, *p* = 0.035), and HVEM on naive CD8+ T cell (OR 0.69, 95% CI: 0.48-0.98, *p* = 0.039); and eight increasing the risk of SSc: Effector Memory CD4-CD8-T cell %T cell (OR 1.85, 95% CI: 1.22-2.82, *p* = 0.004), Effector Memory CD4+ T cell %T cell (OR 1.77, 95% CI: 1.19-2.64, *p* = 0.005), CD4 on HLA DR+ CD4+ T cell (OR 1.76, 95% CI: 1.10-2.80, *p* = 0.018), IgD on IgD+ CD38dim B cell (OR 1.19, 95% CI: 1.02-1.39, *p* = 0.023), IgD on IgD+ CD24- B cell (OR 1.18, 95% CI: 1.02-1.36, *p* = 0.024), IgD on IgD+ CD38+ B cell (OR 1.16, 95% CI: 1.01-1.33, *p* = 0.031), CD45RA- CD4+ T cell Absolute Count (OR 1.41, 95% CI: 1.01-1.95, *p* = 0.042), and CD8 on CD39+ CD8+ T cell (OR 1.35, 95% CI: 1.00-1.81, *p* = 0.046) (Fig 3). MR-PRESSO analysis on these positive exposures indicated no heterogeneous SNPs. Cochran’s Q test (*p* > 0.05) and MR-Egger intercept test (*p* > 0.05) provided strong evidence of no heterogeneity or pleiotropy (Supplementary Table S9). After FDR correction, two immune cells remained significant with *P* values less than 0.05: HLA DR on CD14- CD16+ monocyte (*P*_FDR_ < 0.001) and HLA DR on CD33dim HLA DR+ CD11b+ (*P*_FDR_ < 0.001), indicating a significant causal relationship with SSc. To assess reverse causality, we performed reverse MR analysis with SSc as the exposure and fifteen immune cells as outcomes, all showing no bidirectional causal relationships (*p* > 0.05) (Supplementary Table S11.2).

**Fig 3.**
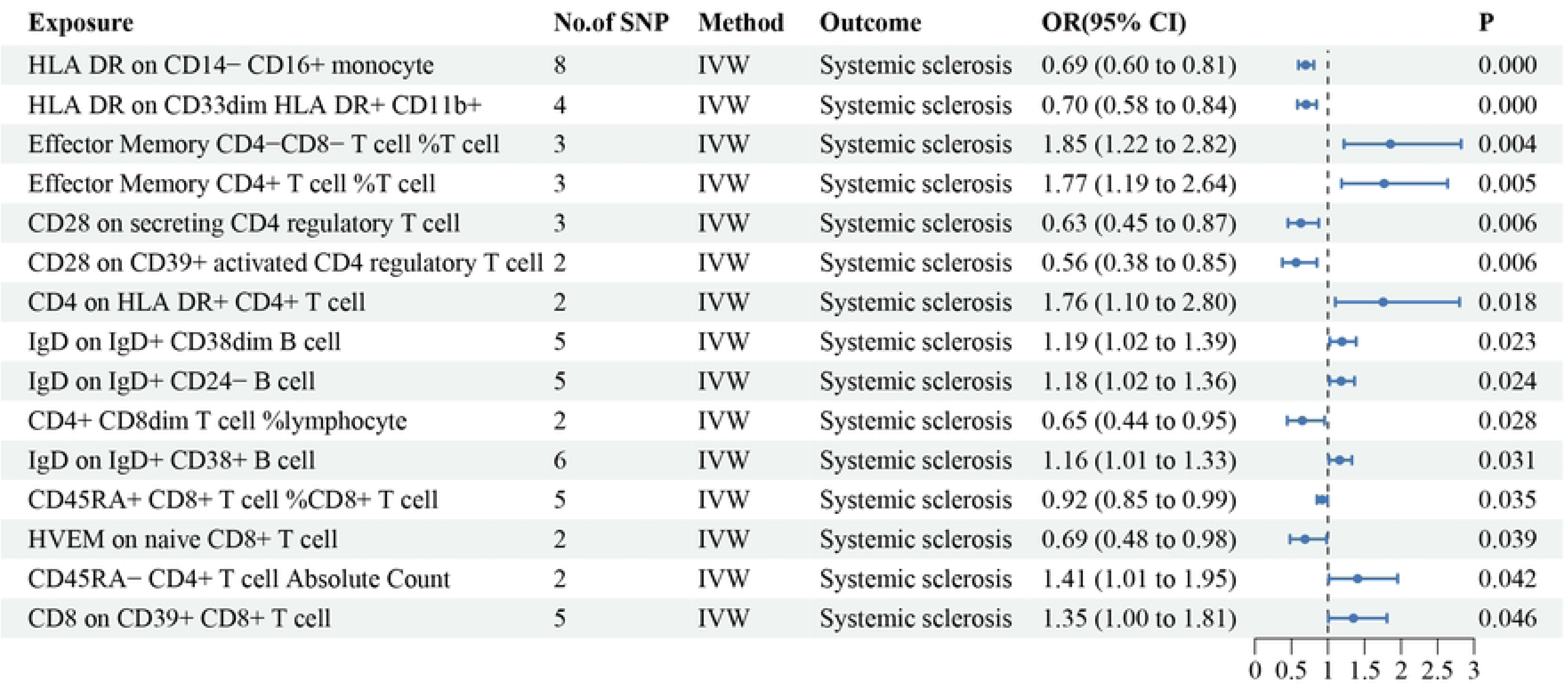

### 3.3 Bidirectional MR Analysis of Blood Metabolites and Metabolite Ratios on SSc

After stringent control of IV quality, the MR study ultimately captured 777 metabolites and 192 metabolite ratios. All included SNPs had an F statistic more remarkable 10 (Supplementary Table S4). Subsequent MR analysis showed significant estimates from IVW (*p* < 0.05). Consistent directions and magnitudes were observed across IVW, MR-Egger, and weighted median estimations. We identified twelve metabolites and two metabolite ratios that may causally influence SSc, including twelve protective factors: Tetradecadienoate (14:2) levels (OR 0.37, 95% CI: 0.18-0.77, *p* = 0.008), X-12844 levels (OR 0.65, 95% CI: 0.47-0.91, *p* = 0.012), 5-acetylamino-6-formylamino-3-methyluracil levels (OR 0.83, 95% CI: 0.70-0.97, *p* = 0.023), Glutamine levels (OR 0.62, 95% CI: 0.40-0.97, *p* = 0.036), Sphingomyelin (d18:1/19:0, d19:1/18:0) levels (OR 0.69, 95% CI: 0.49-0.98, *p* = 0.036), Glycolithocholate sulfate levels (OR 0.62, 95% CI: 0.40-0.97, *p* = 0.037), Octadecanedioylcarnitine (C18-DC) levels (OR 0.80, 95% CI: 0.65-0.99, *p* = 0.038), Gamma-glutamyl-alpha-lysine levels (OR 0.46, 95% CI: 0.22-0.97, *p* = 0.040), 2’-deoxyuridine to cytidine ratio (OR 0.53, 95% CI: 0.29-0.97, *p* = 0.040), X-16935 levels (OR 0.77, 95% CI: 0.59-0.99, *p* = 0.043), Homoarginine levels (OR 0.78, 95% CI: 0.61-0.99, *p* = 0.044), and 1-(1-enyl-palmitoyl)-2-oleoyl-gpc (p-16:0/18:1) levels (OR 0.60, 95% CI: 0.36-1.00, *p* = 0.048); and two risk factors: Paraxanthine to 5-acetylamino-6-formylamino-3-methyluracil ratio (OR 1.18, 95% CI: 1.02-1.36, *p* = 0.029), and 5-hydroxyindole sulfate levels (OR 2.63, 95% CI: 1.02-6.76, *p* = 0.045) ( Fig 4). The presence of heterogeneous SNPs was not supported by MR-PRESSO analysis on these positive exposures (Table S9). Strong evidence was found against heterogeneity and pleiotropy by Cochran’s Q test (*p* > 0.05) and the MR-Egger intercept test (*p* > 0.05) (Supplementary Table S9). After FDR correction, the *P* values for these fourteen metabolites were all greater than 0.05 (Supplementary Table S7), indicating a potential causal relationship with SSc. To assess reverse causality, we performed reverse MR analysis with SSc as the exposure and fourteen metabolites as outcomes, all showing no bidirectional causal relationships (*p* > 0.05) (Supplementary Table 11.3).

**Fig 4.**
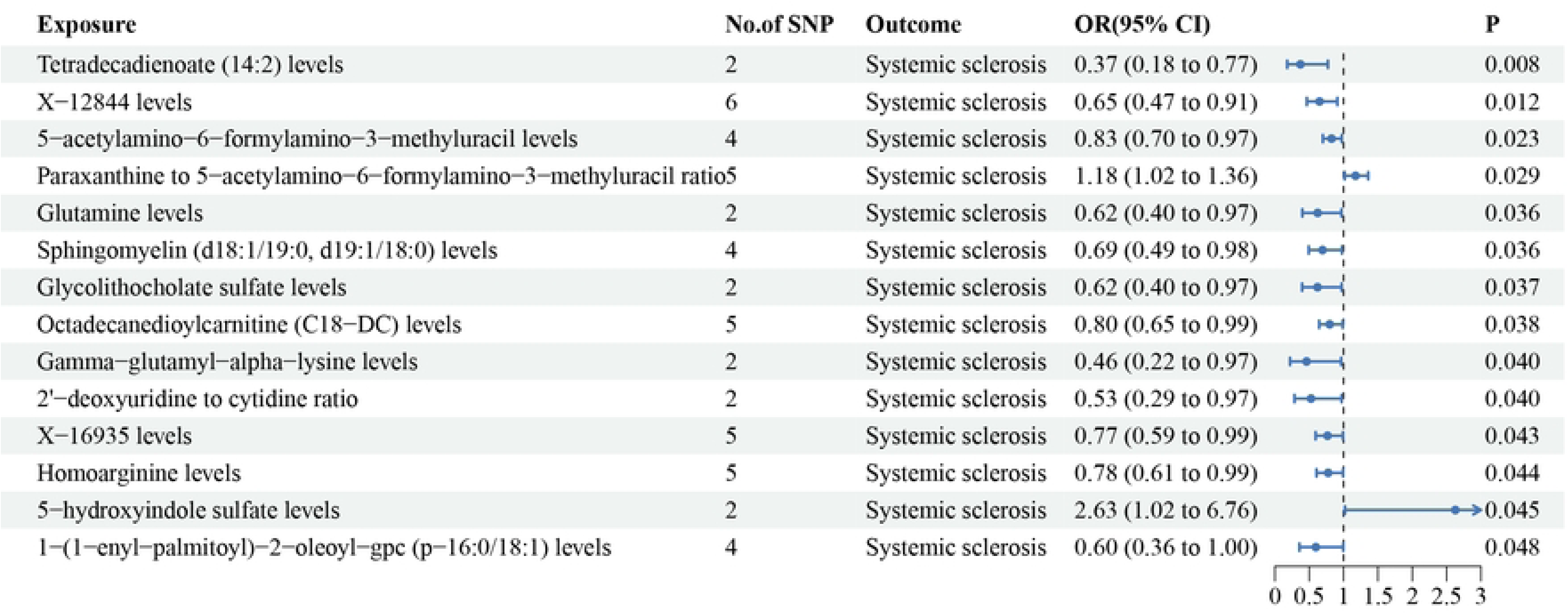

### 3.4 Multivariable MR Results

We used MVMR to further account for the independent causative effects of inflammatory proteins, immunological cells, and metabolites on SSc, considering the intricate interrelationships among linked inflammatory proteins, immune cells, and metabolites. Among the four included inflammatory proteins, two proteins (Hepatocyte growth factor levels, OR 3.377, 95% CI: 1.541-7.401, *p* = 0.002 and Stem cell factor levels, OR 1.386, 95% CI: 1.010-1.902, *p* = 0.043) independently impacted SSc as risk factors. Among the fifteen included immune cells, HLA DR on CD14- CD16+ monocyte (OR 0.380, 95% CI: 0.153-0.943, *p* = 0.037) independently exerted a protective causal effect on SSc. Within the fourteen metabolites, Homoarginine levels and Tetradecadienoate (14:2) levels independently conferred protective causal effects on SSc, whereas 5-hydroxyindole sulfate levels independently acted as a risk factor for SSc (Table 1).

**Table 1.**
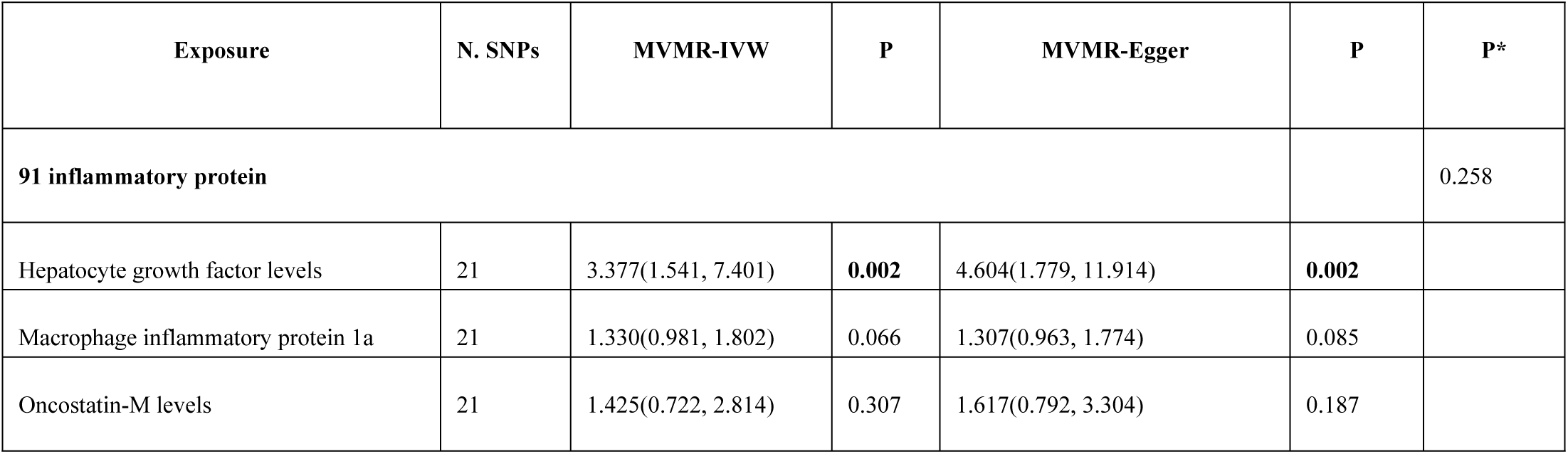
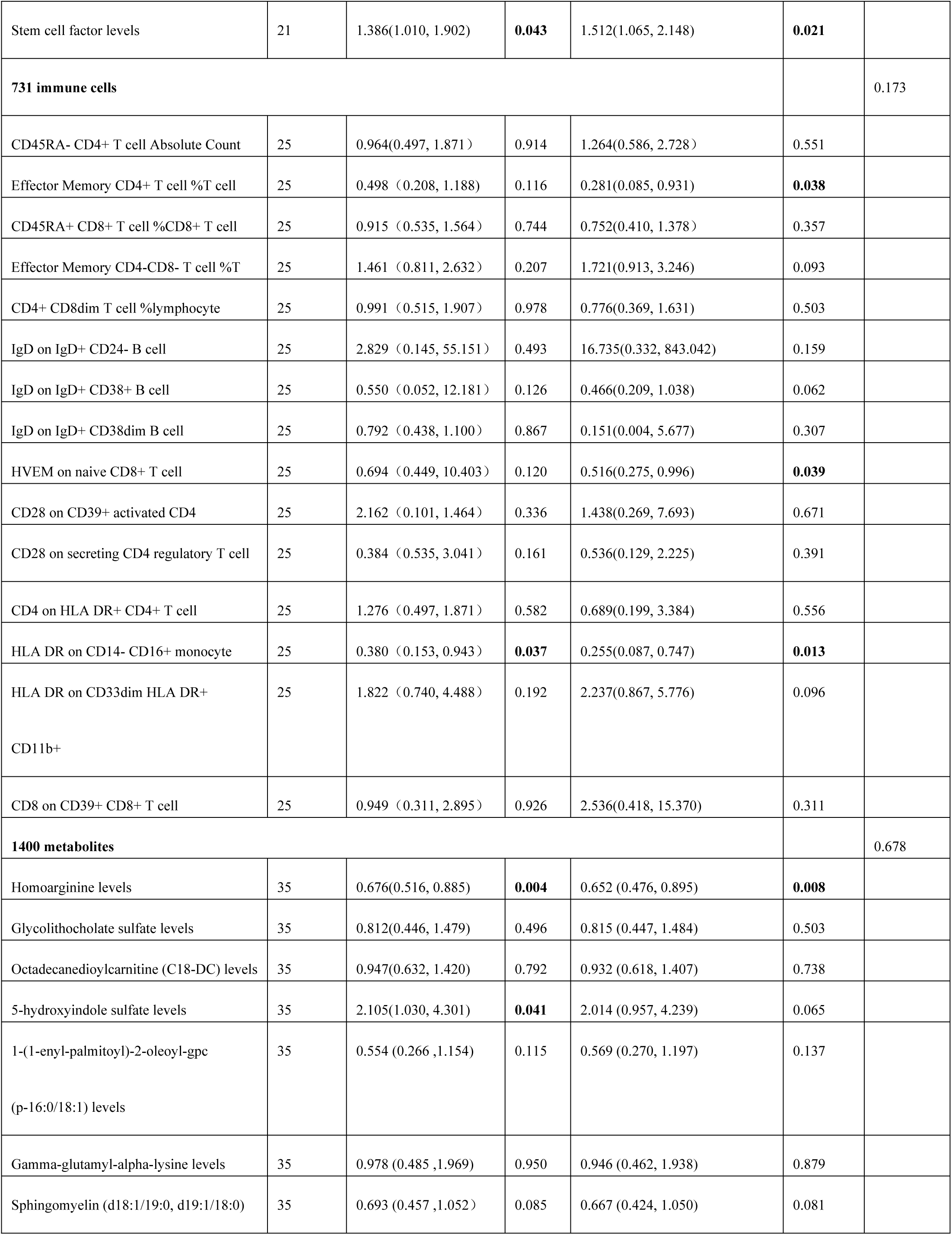
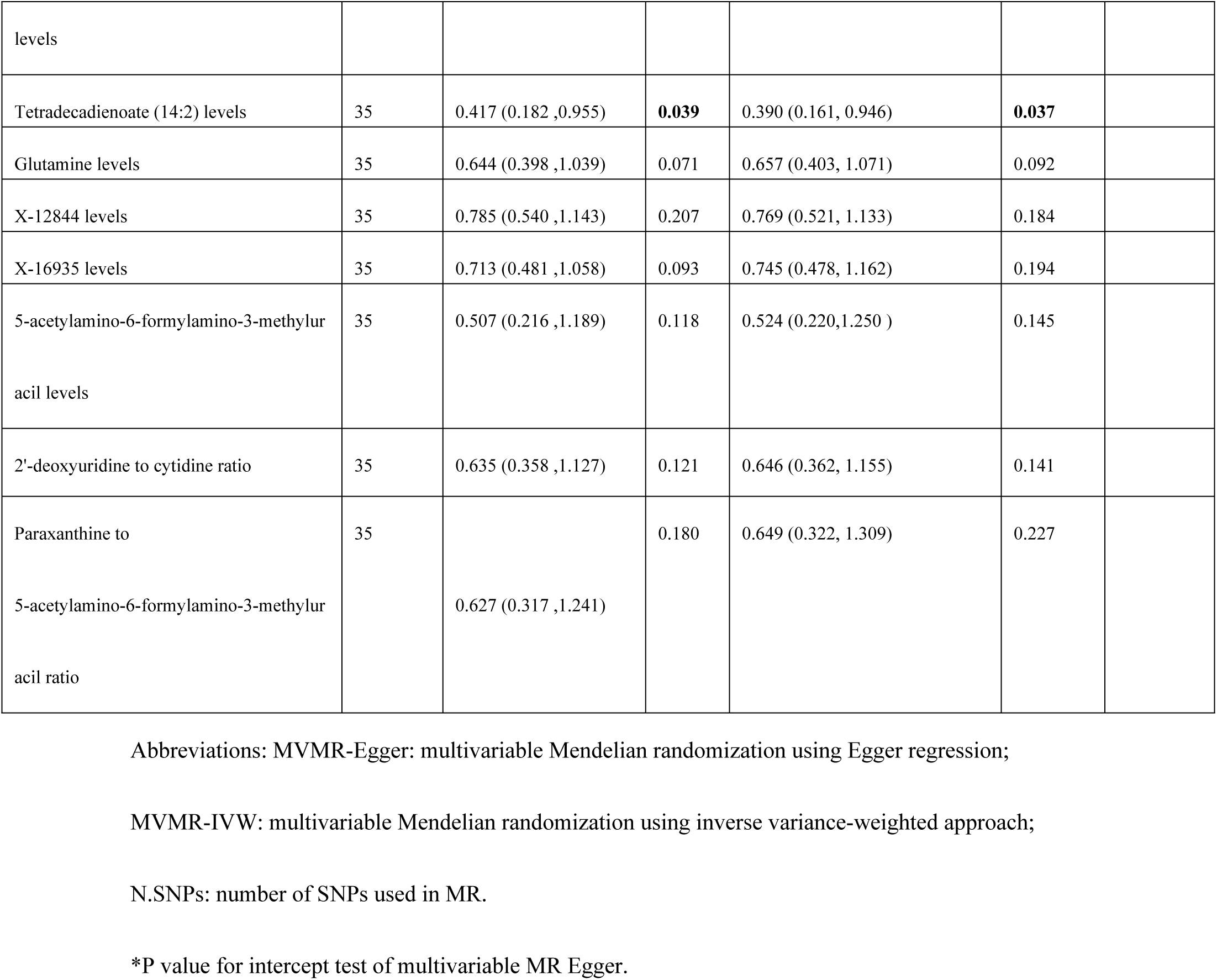
Multivariate Mendelian Randomization Analysis of Identified Inflammatory Proteins, Immune Cells, and Metabolites.

## 4. Discussion

This work is the first to use MR analysis to examine the causal connections between inflammation, metabolism, and SSc. We found a causal association between inflammatory factors and metabolic substances and SSc. Thus, by examining causal linkages and pertinent interactions within the genomic context of SSc, our study is essential in that it attempts to close knowledge gaps regarding the pathophysiology and therapy of SSc.

Our research found that stem cell factor (SCF) and hepatocyte growth factor (HGF) independently contribute to the risk of developing SSc. Studies have identified differential expression of HGF in patients with systemic sclerosis-associated pulmonary arterial hypertension (SSc-PAH), highlighting HGF as a predictive biomarker for SSc-PAH [33]. Elevated HGF was significantly linked to the development of pulmonary arterial hypertension in a prospective cohort study involving 300 SSc patients. This suggests that molecular markers involved in angiogenesis reflect vascular disturbances, with an initial elevation of HGF predictive of a higher likelihood of PAH [34]. This finding is consistent with our results. Interestingly, some studies have suggested the therapeutic benefits of HGF in SSc. HGF increases Matrix Metalloproteinase-1 (MMP-1) levels by activating the transcription factor Ets-1 and inhibits collagen accumulation in SSc fibroblasts by reducing the expression of connective tissue growth factor (CTGF). Similar observations have been made in pulmonary fibroblasts of SSc patients [35]. HGF-modified mesenchymal stem cells (HGF/MSCs) regulate CD4+ T cell polarization and activity, reducing the expression of pro-inflammatory cytokines (IL-1β, IFN-γ, TNF-α, and IL-17A) while increasing anti-inflammatory cytokines (IL-4 and IL-10), thereby mitigating inflammatory responses [36–39]. This contrasts with our study, possibly due to HGF being a potential risk factor for SSc-PAH patients while exerting protective effects in SSc fibrosis and inflammation, suggesting the need for tailored approaches based on the disease environment of individual patients.

SCF, a critical cytokine, accumulates mast cells promoted by SCF derived from keratinocytes in Th17-mediated inflammatory environments [40]. SCF and monocyte chemoattractant protein-1 (MCP-1) induce fibroblast transformation [41]. Studies have also shown significantly higher expression levels of SCF in the epidermis and dermis of SSc patients compared to normal skin, correlating with skin itching, increased mast cell numbers, and pigmentation. This suggests that SCF may be involved in melanocyte activation and the development of dermal mast cells [42]. Furthermore, oncostatin-M (OSM), a member of the IL-6 family, has been implicated in endothelial cell (EC) injury and activation in SSc via OSMRβ and signal transducer and activator of transcription 3 (STAT3), contributing to the development of SSc vascular disease [43]. These findings are consistent with our study results.

Moreover, we confirmed two immune cells causally associated with SSc. HLA-DR on CD14−CD16+ monocytes exhibited the strongest and independent protective effect (*p* = 2.24E-06). Our research indicates that a lower risk of acquiring SSc correlates with increased CD14−CD16+ monocytes. Innate blood cells, known as monocytes, maintain vascular homeostasis and react promptly to pathogens during acute infections. Among these cells are non-classical monocytes known as CD14−CD16+ monocytes, which have anti-inflammatory characteristics and act as the first line of defense against pathogens [44]. Animal studies have shown that non-classical monocytes may enhance regulatory T cell activity through C-X-C motif chemokine ligand 12 (CXCL12) and transforming growth factor-beta (TGF-β) production, thereby controlling inflammation in arthritis models [45]. In lupus nephritis (LN) models, non-classical monocytes accumulate in glomerular vessels and contribute to maintaining glomerular homeostasis [46]. CD14−CD16+ monocytes have been shown to exhibit anti-inflammatory, vascular homeostasis, and cardiovascular protective effects, demonstrating benefits in controlling autoimmune diseases [45–47]. Our study also identified a protective role of CD14−CD16+ monocytes in SSc, suggesting their therapeutic potential in autoimmune diseases.

In addition to HLA-DR on CD14−CD16+ monocytes, we should also focus on effector memory CD4+ T cells, which may increase the risk of SSc development. CCR7−CD4+ memory T cells and effector cells produce intracellular TNFα, IL-13, and IL-4, particularly in diffuse cutaneous systemic sclerosis (dcSSc). SSc-derived CD4+ T cells show an increased proportion of CCR7− memory T cells following inadequate Treg suppression, highlighting specific CD4+ memory T cell subsets that significantly promote pro-inflammatory cytokine production in dcSSc [48]. Our study underscores the crucial role of T cell memory phenotypes in controlling inflammation in SSc patients, providing direction for further research.

We established potential causal relationships between 14 metabolites and SSc, with 13 as protective factors. Notably, multivariate analysis identified 5-hydroxyindole sulfate as an independent risk factor. These metabolites, which are involved in central metabolic pathways in patients with SSc, are mainly composed of fatty acids, amino acids, glycerophospholipids, and bile acids. According to the study, the principal pathways impacted by skin involvement are steroid hormone biosynthesis and amino acid metabolism, while glutamine metabolism is the primary process changed in SSc patients with lung involvement [49]. Homoarginine has been a controversial factor in vascular disease. However, recent research indicates it is a protective and prognostic factor in cardiovascular disease, reducing atherosclerosis and serving as a therapeutic target [50]. Supplementing homoarginine in apolipoprotein E-deficient mice regulates the T-cell cytoskeleton and inhibits pseudopodia formation by myosin heavy chain 9 (Myh9), affecting T-cell migration and proliferation, thereby alleviating atherosclerosis [51]. Homoarginine supplementation significantly improved albuminuria, renal inflammation, and oxidative stress in diabetic mice, showing potential therapeutic benefits in diabetic nephropathy [52]. Tetradecadienoate (14:2) may play a role in inflammatory processes; while excess ω-6 fatty acids may increase the risk of inflammatory diseases, appropriate proportions help maintain cardiovascular health [53]. Currently, no studies have linked tetradecadienoate (14:2) to SSc. Our findings suggest that homoarginine and tetradecadienoate (14:2) may benefit vascular lesions in SSc.

5-Hydroxyindole Sulfate (5-HIAA) is a 5-Hydroxytryptamine (5-HT; serotonin) metabolite that causes pulmonary vascular resistance, stimulates vasoconstriction, encourages platelet aggregation, and causes skin and visceral organ fibrosis [54]. Researchers have reported two cases where the 5-HT3 receptor antagonist tropisetron successfully treated rheumatoid arthritis (RA) and showed efficacy in refractory rheumatoid vasculitis, highlighting the need for more extensive clinical studies to validate these findings further [55]. 5-HT activates the 5-HT_2B_ receptor in TGF-β-dependent manner, significantly inducing extracellular matrix synthesis in interstitial fibroblasts. Skin fibrosis was alleviated in mice lacking the 5-HT_2B_ gene, suggesting that the 5-HT/5-HT_2B_ signaling pathway may represent a novel therapeutic target for fibrotic diseases [56]. Further studies have found that 5-HT inhibitors may reduce fibrosis by inhibiting the non-canonical signaling pathway mediated by TGF-β1 [57]. These observations hold significant therapeutic implications for fibrotic diseases, including SSc. Based on our research findings, we speculate that 5-HT inhibitors could serve as potential therapeutic targets for SSc.

By utilizing the most significant sample size from GWAS cohorts, our work resolved disagreements seen in observational research by thoroughly examining the connections and causal links between inflammatory proteins, immune cells, metabolites, and SSc. However, it is vital to recognize a few of our research’s shortcomings. First, the fact that only people of European ancestry were included in the GWAS summary data limits the applicability of our findings to populations with diverse genetic origins. Establishing causality requires taking ethnicity into account. Second, we could not obtain sample data from other organs, such as the skin or lungs, and instead confined our analysis to the exposure levels in peripheral blood. Lastly, The current GWAS data do not include individual patient-level information, precluding further subgroup analysis within the existing SSc patient population to obtain more precise results.

## 5. Conclusions

Our investigation found four inflammatory proteins, fifteen immune cell characteristics, fourteen metabolites, and SSc causally related. These results resolve disagreements noted in earlier observational studies and offer fresh perspectives on the pathophysiology of SSc, which may help formulate treatment plans.

## Data Availability

If the data are all contained within the manuscript and/or Supporting Information files, enter the following: All relevant data are within the manuscript and its Supporting Information files.

## Supporting information

**S1 File.**

(DOCX)

**S2 File.**

(XLXS)

## Abbreviations

SSc: Systemic sclerosis
MR: Mendelian Randomization
IL-6: Interleukin-6
SNPs: Single nucleotide polymorphisms
IVs: Instrumental variables
GWAS: Genome-wide association studies
IVW: Inverse variance weighted
LD: Linkage disequilibrium
OR: Odds ratio
CI: Confidence interval
MRPRESSO: MR pleiotropy residual and outlier
FDR: False discovery rate
MVMR: Multivariable Mendelian randomization
SCF: Stem cell factor
HGF: Hepatocyte growth factor
PAH: Pulmonary arterial hypertension
MMP-1: Matrix Metalloproteinase-1
CTGF: Connective tissue growth factor
MSCs: Modified mesenchymal stem cells
MCP-1: Monocyte chemoattractant protein-1
OSM: Oncostatin-M
EC: Endothelial cell
STAT3: Signal transducer and activator of transcription 3
CXCL12: C-X-C motif chemokine ligand 12
TGF-β: Transforming growth factor-beta
LN: Lupus nephritis
dcSSc: Diffuse cutaneous systemic sclerosis
Myh9: Myosin heavy chain 9
5-HIAA: 5-Hydroxyindole Sulfate
RA: Rheumatoid arthritis
5-HT: 5-Hydroxytryptamine

## Declarations

## Ethics approval and consent to participate

Not applicable.

## Clinical trial number

Not applicable.

## Consent for publication

Not applicable.

## Availability of data and materials

All data generated or analysed during this study are included in this published article.

## Authors’ contributions

YZ, WZ, and LL conceptualized the idea for the manuscript. YZ, XY, and JP contributed to the methods for the paper. YZ, ZL, and WZ drafted the manuscript under the supervision of LL and XY. All authors read and approved the final manuscript.

## Acknowledgements

The authors would like to thank the FinnGen database, the public catalog of GWAS, and the Department of Public Health and Primary Care for providing the data used in this study.

